# A predictive atlas of disease onset from retinal fundus photographs

**DOI:** 10.1101/2024.03.15.24304339

**Authors:** Thore Buergel, Lukas Loock, Jakob Steinfeldt, Laura Hoffmann, Steffen Emil Künzel, Julius Upmeier zu Belzen, Anthony P. Khawaja, Robert Luben, Paul J. Foster, Claudia Langenberg, Ulf Landmesser, John Deanfield, Oliver Zeitz, Antonia Joussen, Maik Pietzner, Benjamin Wild, Roland Eils

## Abstract

Early detection of high-risk individuals is crucial for healthcare systems to cope with changing demographics and an ever-increasing patient population. Images of the retinal fundus are a non-invasive, low-cost examination routinely collected and potentially scalable beyond ophthalmology. Prior work demonstrated the potential of retinal images for risk assessment for common cardiometabolic diseases, but it remains unclear whether this potential extends to a broader range of human diseases. Here, we extended a retinal foundation model (RETFound) to systematically explore the predictive potential of retinal images as a low-cost screening strategy for disease onset across >750 incident diseases in >60,000 individuals. For more than a third (n=308) of the diseases, we demonstrated improved discriminative performance compared to readily available patient characteristics. This included 281 diseases outside of ophthalmology, such as type 2 diabetes (Delta C-Index: UK Biobank +0.073 (0.068, 0.079)) or chronic obstructive pulmonary disease (Delta C-Index: UK Biobank +0.047 (0.039, 0.054)), showcasing the potential of retinal images to complement screening strategies more widely. Moreover, we externally validated these findings in 7,248 individuals from the EPIC-Norfolk Eye Study. Notably, retinal information did not improve the prediction for the onset of cardiovascular diseases compared to established primary prevention scores, demonstrating the need for rigorous benchmarking and disease-agnostic efforts to design cost-efficient screening strategies to improve population health. We demonstrated that predictive improvements were attributable to retinal vascularisation patterns and less obvious features, such as eye colour or lens morphology, by extracting image attributions from risk models and performing genome-wide association studies, respectively. Genetic findings further highlighted commonalities between eye-derived risk estimates and complex disorders, including novel loci, such as *IMAP1*, for iron homeostasis. In conclusion, we present the first comprehensive evaluation of predictive information derived from retinal fundus photographs, illustrating the potential and limitations of easily accessible and low-cost retinal images for risk assessment across common and rare diseases.

**Research in context:** *Evidence before this study:* Before undertaking this study, we reviewed the literature on the predictive utility of medical imaging for disease onset, focusing particularly on retinal fundus photographs. We conducted searches in databases including PubMed and Google Scholar, spanning from the inception of these databases to January 1, 2023. Our search terms included “retinal fundus photography”, “disease prediction”, “machine learning”, “deep learning”, and “healthcare AI”, without language restrictions. Prior research has shown the promise of retinal images in diagnosing and predicting a range of conditions, notably within ophthalmology and specific systemic diseases such as diabetes and cardiovascular diseases. However, a comprehensive evaluation of retinal images’ predictive potential across a broad spectrum of diseases, particularly those without known associations to retinal changes, was lacking. Studies identified varied in quality, with many focusing on single diseases or small datasets, indicating a potential risk of bias and overfitting.

*Added value of this study:* Our study extends the application of retinal fundus photographs from ophthalmological and systemic diseases to more than 750 incident diseases, leveraging a foundation model combined with a deep multi-task neural network. This represents the first systematic exploration of the predictive potential of retinal images across the human phenome, significantly expanding the scope of diseases for which these images could serve as a low-cost screening strategy. Moreover, we rigorously compare the predictive value of retinal images against established primary prevention scores for cardiovascular diseases, showing both the strengths and limitations of this approach. This dual focus provides a nuanced understanding of where retinal imaging can complement existing screening strategies and where it may not offer additional predictive value.

*Implications of all the available evidence:* The evidence from our study, combined with existing research, suggests that retinal fundus photographs hold promise for predicting disease onset across a wide range of conditions, far beyond their current use. However, our work also emphasizes the importance of contextualizing these findings within the broader landscape of available prediction tools and established primary prevention. The implications for practice include the potential integration of retinal imaging into broader screening programs, particularly for diseases where predictive gains over existing methods are demonstrated. For policy, our findings advocate for further investment in AI and machine learning research in healthcare, particularly in methods that improve upon or complement existing prediction models. Future research should focus on refining these predictive models, exploring the integration of retinal imaging with other biomarkers, and conducting prospective studies to validate the clinical utility of these approaches in diverse populations.

## Introduction

The progressive change in demographics in most industrialised countries leading to an ever-increasing number of patients with multiple diseases requiring hospitalisation, but also the insufficient health care capabilities in most low- and middle-income countries require a transformative change in how we identify individuals at high risk early to avoid or delay severe disease onset. However, it is currently largely unclear what low-cost, scalable approaches exist to achieve this aim at population scale for possibly many diseases simultaneously.

Retinal fundoscopy is a non-invasive, simple, and fast procedure allowing high-resolution imaging of the retina. Retinal features, such as vascularisation patterns, have already been associated with patients’ vascular^2,3^ and neurological conditions^4,5^ and often manifest subclinically years before the onset of diseases^4,6–9^. Computer vision techniques have been used to extract features that predict demographic factors, such as age and biological sex^10,11^, and a wide range of vascular and systemic measures^10,12–14^, such as the glomerular filtration rate^15^, coronary artery calcium scores^16^ or lipid profiles^12^. Additionally, models have been developed to diagnose or predict the onset of specific diseases using retinal fundus photographs, including age-related macular degeneration^17–19^, diabetic retinopathy^20,21^, anemia^22^, chronic kidney disease^15,23^, type 2 diabetes^15^, major adverse cardiac events^10^, stroke and myocardial infarction^24^, Alzheimer’s^25^ as well as all-cause mortality^11,26^. However, a comprehensive evaluation of predictive information derived from retinal fundus photographs and the extent of the information beyond established predictors has not been systematically investigated to date.

Here, we provide a rigorous evaluation of the predictive information in retinal fundus photographs across the human phenome. We simultaneously learned disease-specific risk estimates for 752 diseases in the UK Biobank cohort^28,29^ (Figure 1) by combining a recent foundation model for retinal fundus photographs^27^ with a deep multi-task neural network for disease onset prediction. Our model stratifies the risk of future disease for 542 of these endpoints, most notably for type 2 diabetes, chronic kidney disease, and COPD. We then combined these predicted risks with basic patient characteristics and observed improved predictive performance for more than a third of the diseases tested spanning almost all clinical specialties, including diseases without known retinal associations, such as chronic obstructive pulmonary disease, heart failure, a subset of malignant neoplasms, and mental disorders, such as major depression. These findings exemplify the potential clinical utility of retinal images for risk stratification. Moreover, we externally validated these findings in 7,248 individuals from the EPIC-Norfolk Eye Study^31,32^. Finally, we demonstrate the explainability of our models by identifying genetic risk loci for informative risk scores and extracting biologically plausible features, like vascularisation patterns, using image attribution techniques.

**Figure 1:**
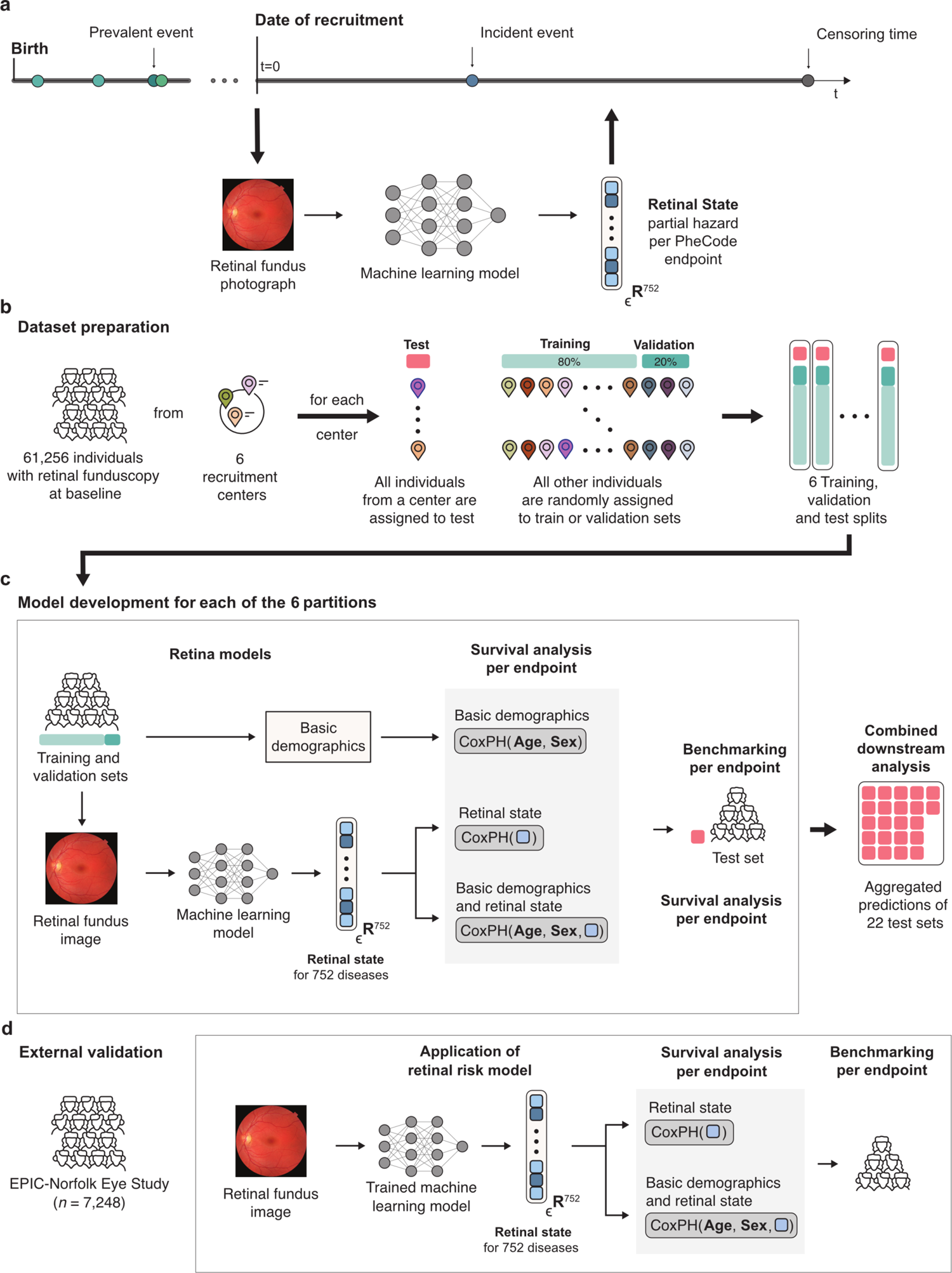
Overview of the study. **a)** Retinal fundus photographs are taken at recruitment. Here, we build upon a foundation model for retinal images and extend it to predict phenome-wide incident disease onset for 752 endpoints **b)** to estimate retinal states from retinal fundus photographs. The eligible UK Biobank population (with retinal fundoscopy at baseline and valid consent) was split into train, validation and test sets in six-fold nested cross-validation based on the assigned UK Biobank assessment centre where eye tests were carried out. **c)** The Retinal State Model was trained on fundus photographs to predict endpoint-specific retinal risks against 752 phecode endpoints for each of the six partitions. Subsequently, for each endpoint, Cox Proportional Hazard (CPH) Models were developed on the metabolomic state in combination with sets of commonly available clinical predictors to model disease risk. Predictions of the CPH model on the test set were aggregated for downstream analysis. **d)** The models were externally validated in the independent EPIC-Norfolk cohort. Models were transferred and used without retraining before extracting the retinal states.

## Results

### Characteristics of the study populations and the retinal risk model

In this study, we rigorously investigated the predictive information of the human retina using advanced machine learning methods, evaluating its potential as a low-cost screening tool. To that end, we derived integrated retinal states capturing information on incident disease risk in a sample of the general population in the UK Biobank cohort^28,29^ (Figure 1a,b, Table 1). The study population, represented in Table 1, included 61,256 individuals who underwent retinal fundoscopy upon recruitment. The subcohort with retinal images was representative of the study population. The median follow-up time was 11.4 years, accounting for approximately 685,000 person-years of follow-up.

To extract the retinal risk estimates, we used a multi-task vision transformer network based on the pre-trained RETFound foundation model^27^ fine-tuned on retinal fundus photographs to simultaneously model the integrative retinal state of 752 endpoints throughout the entire human phenome (Figure 1a, Supplementary Figure 1, Supplementary Table 1, Methods Section “Retinal risk model”). The model allowed us to systematically extract relevant information from retinal fundus photographs while maintaining the flexibility to fit endpoint-specific variations. Subsequently, we used the extracted retinal risk estimates in Cox Proportional Hazard models to predict disease onset in a spatial cross-validation scheme.

We assessed the predictive value of these risk estimates both independently and in conjunction with readily available demographic factors, such as age and biological sex, and other potential predictors (Supplementary Table 2). Subsequently, we collected all estimates for the 752 endpoints to create a phenome-wide atlas of predictive information from retinal fundus photographs (Fig 1c). We externally validated the atlas of predictive retinal information by applying our developed retinal state models to retinal fundus photographs of the EPIC-Norfolk Eye Study^33,34^ (Fig 1d; Supplementary Table 8).

### Retinal information stratifies phenome-wide incident disease onset

We initially focused on identifying associations between retinal fundus images and the likelihood of future disease onset. We examined how retinal features identified by our model without adjusting for any other factors correlate with the risk of developing 752 different diseases across the human phenome (Figure 2a). To that end, we measured the incidence of diseases in relation to the calculated risk levels from retinal images and compared the disease occurrence rates between individuals in the highest and lowest 10% of risk scores. As an illustration, the risk stratification for all-cause mortality is depicted in Figure 2b, while aggregated metrics across diseases are presented in Figure 2c.

**Figure 2:**
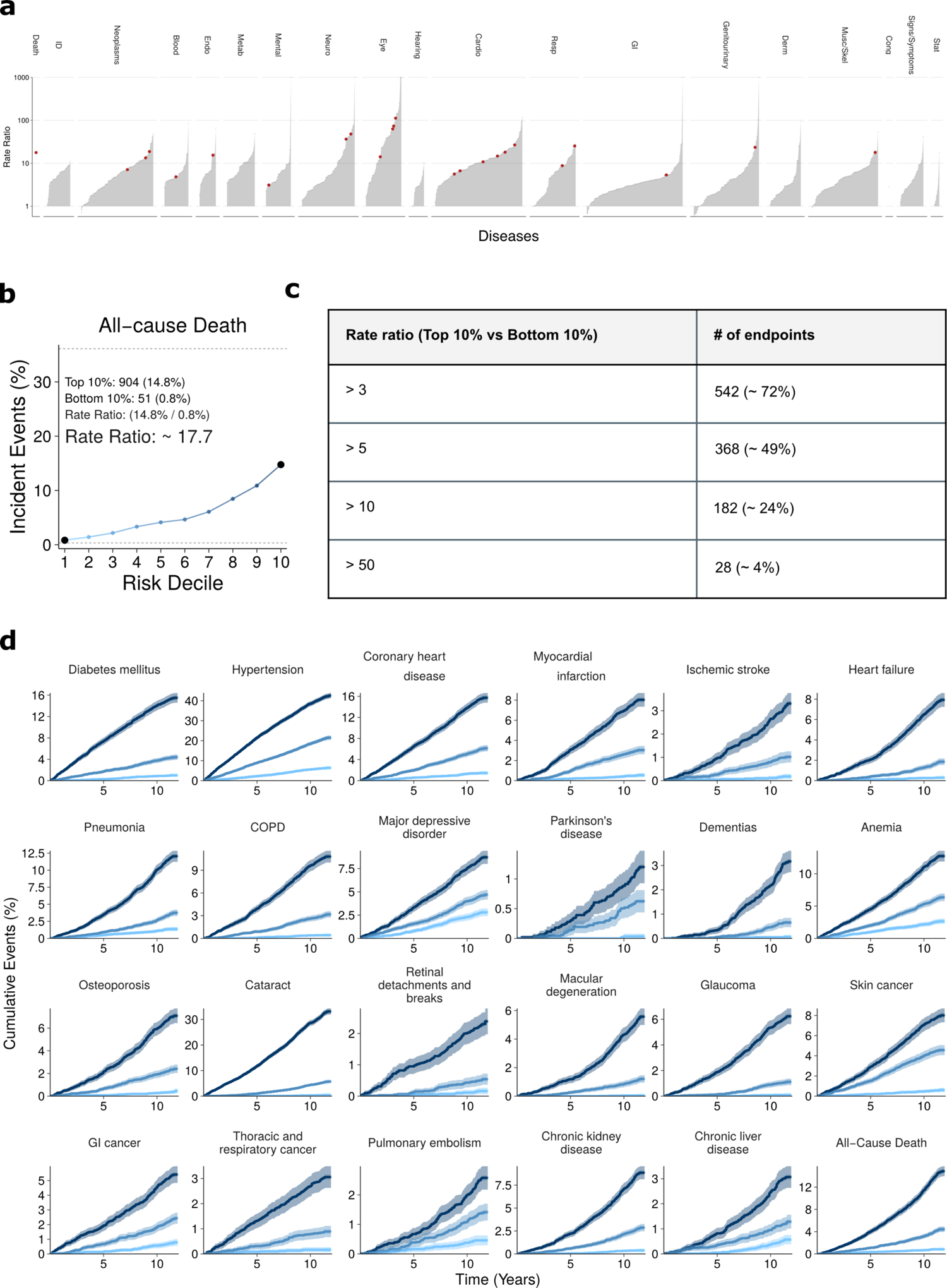
Retinal information stratifies phenome-wide incident disease onset. **a)** Ratio of incident events in the Top 10% compared with the Bottom 10% of the estimated risk estimates. Red dots indicate the selected endpoints displayed in Fig 2d. b) To illustrate, 904 (14.8%) individuals in the top risk decile for all-cause death experienced an event compared with only 51 (1%) in the bottom decile, with a rate ratio of 17.7. c) Overview of the number of endpoints surpassing a given rate ratio. d) Cumulative event rates for a selection of 24 endpoints for the Top 10%, median, and Bottom 10% of risk percentiles over 15 years. Statistical measures were derived from 61,256 individuals. Individuals with prevalent diseases were excluded from the endpoints-specific analysis.

We observed monotonous increases in the event rates with increasing risk state percentiles, indicating a stratification of high and low-risk individuals for most endpoints, covering a broad range of disease categories and etiologies. (Figure 2a,c): For 542 out of the 752 endpoints (∼72%; Supplementary Table 3), we observed more than three times as many events for individuals in the top 10% of predicted risk estimates compared to those in the bottom 10%, with risk ratios reaching as high as 80 for Alzheimer’s diseases (Ratio ∼ 80.0), and spanning a diverse set of diseases like type 2 diabetes (Ratio ∼ 16.5), chronic kidney disease (Ratio ∼ 23.4), coronary heart disease (Ratio ∼ 10.8), lung cancer (Ratio ∼ 28.5), or Parkinson’s disease (Ratio ∼ 36.5). These results largely replicated in the EPIC-Norfolk cohort, where we evaluated our retinal state model on 173 endpoints and identified rate ratios greater than 1 for 165 (95%) of the 173 endpoints examined, with 114 (66%) having rate ratios greater than 3, 73 (42%) greater than 5, 31 (18%) greater than 10, and 3 (2%) greater than 50 (Supp. Fig 2a, Supplementary Table 3).

We then selected 24 endpoints encompassing common diseases across various clinical specialties and diseases with established retinal associations for further analysis. We present cumulative event rates for these endpoints for up to 15 years of follow-up, categorised into top, median, and bottom percentiles (Fig 2d). For 17 of these 24 endpoints with sufficient sample size, we validated these findings in the EPIC-Norfolk cohort and found that the observed associations were largely consistent across different cohorts (Suppl Figure 2, Supp. Table 3).

### Retinal information improves discriminative performance for a subset of diseases

Retinal fundus photographs significantly improved the predictive performance compared to a baseline based on age and biological sex for 27 (71%) out of the 38 investigated eye-related diseases (using Cox Proportial Hazards (CPH) models, see Methods for details). These endpoints include cataracts (C-Index: 0.74 (CI 0.73, 0.74) → 0.80 (CI 0.77, 0.80)), macular degeneration (C-Index: 0.74 (CI 0.73, 0.75) → 0.79 (CI 0.77, 0.79)), retinal detachments and breaks (C-Index: 0.60 (CI 0.57, 0.62) → 0.73 (CI 0.71, 0.75)), and myopia (C-Index: 0.61 (CI 0.59, 0.63) → 0.87 (CI 0.86, 0.88)) (Figure 3b).

**Figure 3:**
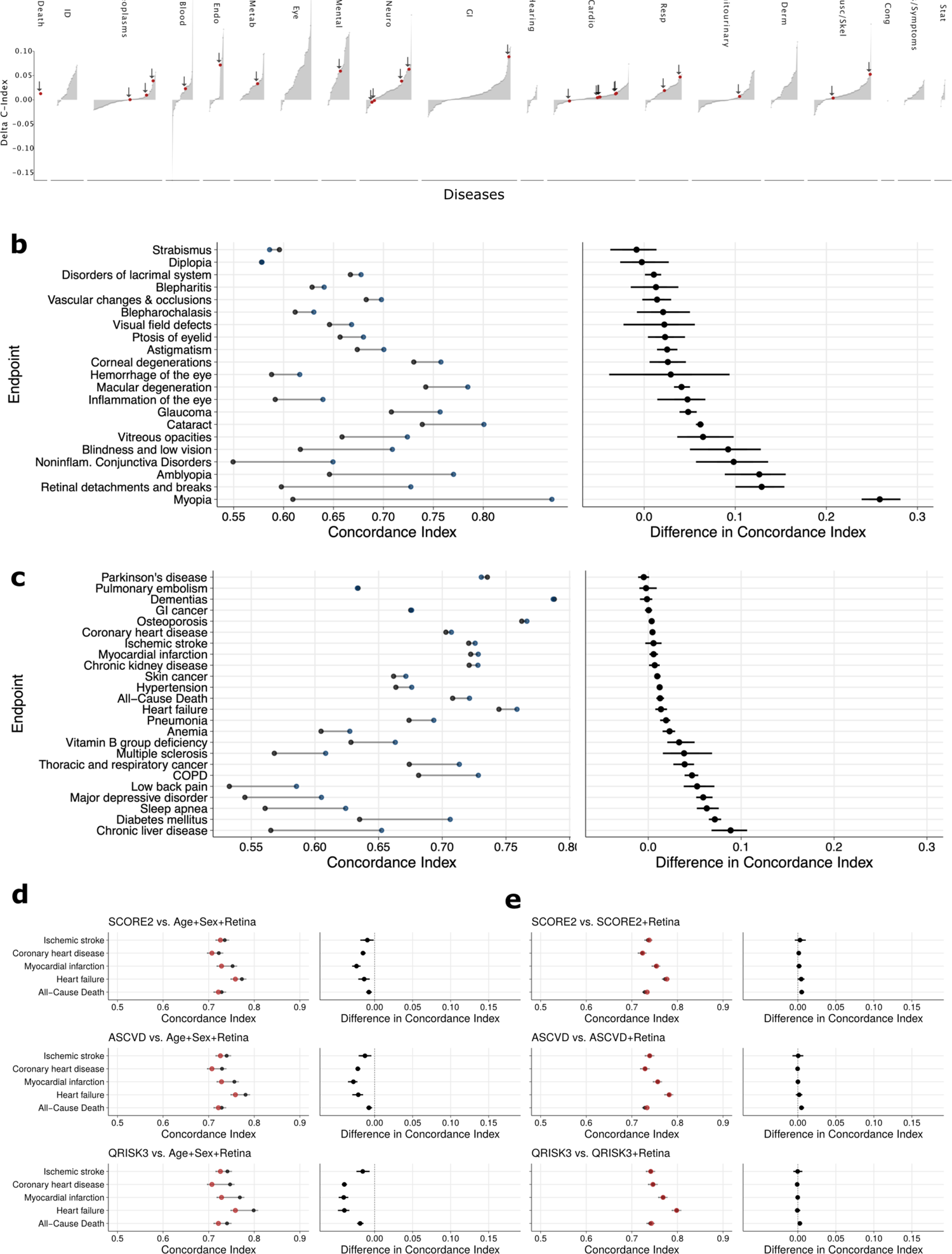
Retinal information improves discriminative performance for a subset of diseases. **a)** Differences in discriminatory performance quantified by the C-Index between CPH models trained on Age+Sex and Age+Sex+Retina for all 752 endpoints. Significant improvements over the baseline model (Age+Sex, age, and biological sex only) are found for 308 (40.96%) of the 752 investigated endpoints. Red dots indicate selected endpoints in Fig. 3b. **b-c)** Absolute discriminatory performance in terms of C-Index comparing the baseline (Age+Sex, black point) with the added retinal information (Age+Sex+Retina, red point) for selected ophthalmological **(b)** and diverse **(c)** endpoints. **d)** Discriminatory performances (left) and direct differences (right) in terms of C-index between Cox Proportional Hazards (CPH) models trained on sets of established cardiovascular predictors (SCORE2, ASCVD, and QRISK3 as black dots) and their extensions with retinal information (red dots). **e)** Discriminatory performances (left) and direct differences (right) in terms of C-index between CPH models trained on sets of established cardiovascular predictors (SCORE2, ASCVD, and QRISK3 as black dots) and a simplified risk model based on Age+Sex+Retina. Dots indicate medians and whiskers extend to the 95% confidence interval for a distribution bootstrapped over 100 iterations.

We observed improved predictive performance for 308 (40.96%) of the 752 investigated endpoints, covering a broad range of etiologies (Figure 3, Supplementary Table 4). This included conditions without established retinal associations, notably chronic obstructive pulmonary disease (C-Index: 0.68 (CI 0.67, 0.69) → 0.73 (CI 0.72, 0.74)), heart failure (C-Index: 0.74 (CI 0.74, 0.75) → 0.76 (CI 0.76, 0.77)), malignant neoplasms of thoracic and respiratory organs (C-Index: 0.67 (CI 0.66, 0.69) → 0.71 (CI 0.70, 0.73)), and mental disorders like major depression (C-Index: 0.55 (CI 0.54, 0.55) → 0.61 (CI 0.60, 0.61)). We observed improved predictive performance for all-cause mortality (C-Index: 0.71 (CI 0.70, 0.72) → 0.72 (CI 0.72, 0.73)). We also found significant predictive improvements with potential clinical utility for diseases such as chronic liver disease (C-Index: 0.57 (CI 0.55, 0.58) → 0.65 (CI 0.64, 0.67)), type 2 diabetes (C-Index: 0.64 (CI 0.63, 0.64) → 0.71 (CI 0.71, 0.72)), and multiple sclerosis (C-Index: 0.57 (CI 0.53, 0.60) → 0.61 (CI 0.57, 0.64)). Notably, we found that 83.3% of the metabolic, 74.1% of the mental, and 71.1% of all assessed eye-related endpoints, but only 29.3% of the cardiovascular endpoints and 11.8% of the neoplasms benefited from including retinal information in our model (Supplementary Table 5). The adjusted hazard ratios (HR, per SD retinal state, with 95% CI) of the CPH models augmented with the retinal information are presented in Supplementary Table 6.

We observed a slightly lower (31.8%, 55 out of 173 diseases with superior performance) validation rate of our model in the independent EPIC-Norfolk cohort (Supplementary Figure 4a, Supplementary Table 4). However, missing replication for endpoints like all-cause death indicates that different cohort characteristics (people in EPIC-N were, on average, 9 years older and less likely to be current smokers, see Supplementary Tables 1 and 8) potentially require retraining of the final models.

Apart from eye disorders, retinal images have repeatedly been proposed for the risk stratification of cardiovascular diseases^10^. However, we did not observe significant improvements when rigorously benchmarking retinal risk estimates with established clinical risk scores (e.g., European Society of Cardiology SCORE2^35^, the American Heart Association ASCVD^36^, and the British QRISK3^37^ score, recommended by the NHS Health Check^38^) (Figure 3c). In addition, augmenting the SCORE2, ASCVD, and QRISK3 with the retinal information did not yield significant discriminatory improvements for any cardiovascular endpoint (see Figure 3d and Supplementary Table 7), suggesting that retinal images capture information already in clinical use for risk stratification.

### Image attributions characterise the predictive information in the retina

To localise retinal fundus regions linked with disease risk, we used information bottleneck attributions (IBA)^41^ for six prevalent diseases characterised by high morbidity burdens (Figure 4, Supplementary Figure 4; refer to Methods for further details). The resulting attribution maps demonstrated that the model extracts information from the retinal fundus background and specific landmarks, such as vascular patterns, the fovea, and the macula. Notably, we observed significant differences in risk attributions when compared to a model that is explicitly adjusted for age and sex, thus highlighting the predictive information that is additive to these basic demographics (Supplementary Figure 4, see Methods for details). Interestingly, the attributed regions often remained consistent for an individual across different endpoints (Figure 4), indicating that general individual-level features, rather than disease-specific ones, play a key role in mediating the information on multi-disease risk in the human retina. Understanding the model is imperative for successful clinical application^39,40^. Our attribution method evaluates information that both increases or decreases predicted risk, enabling the creation of attribution maps that delineate this risk along with factors beyond age and sex (Supplementary Figure 4).

**Figure 4:**
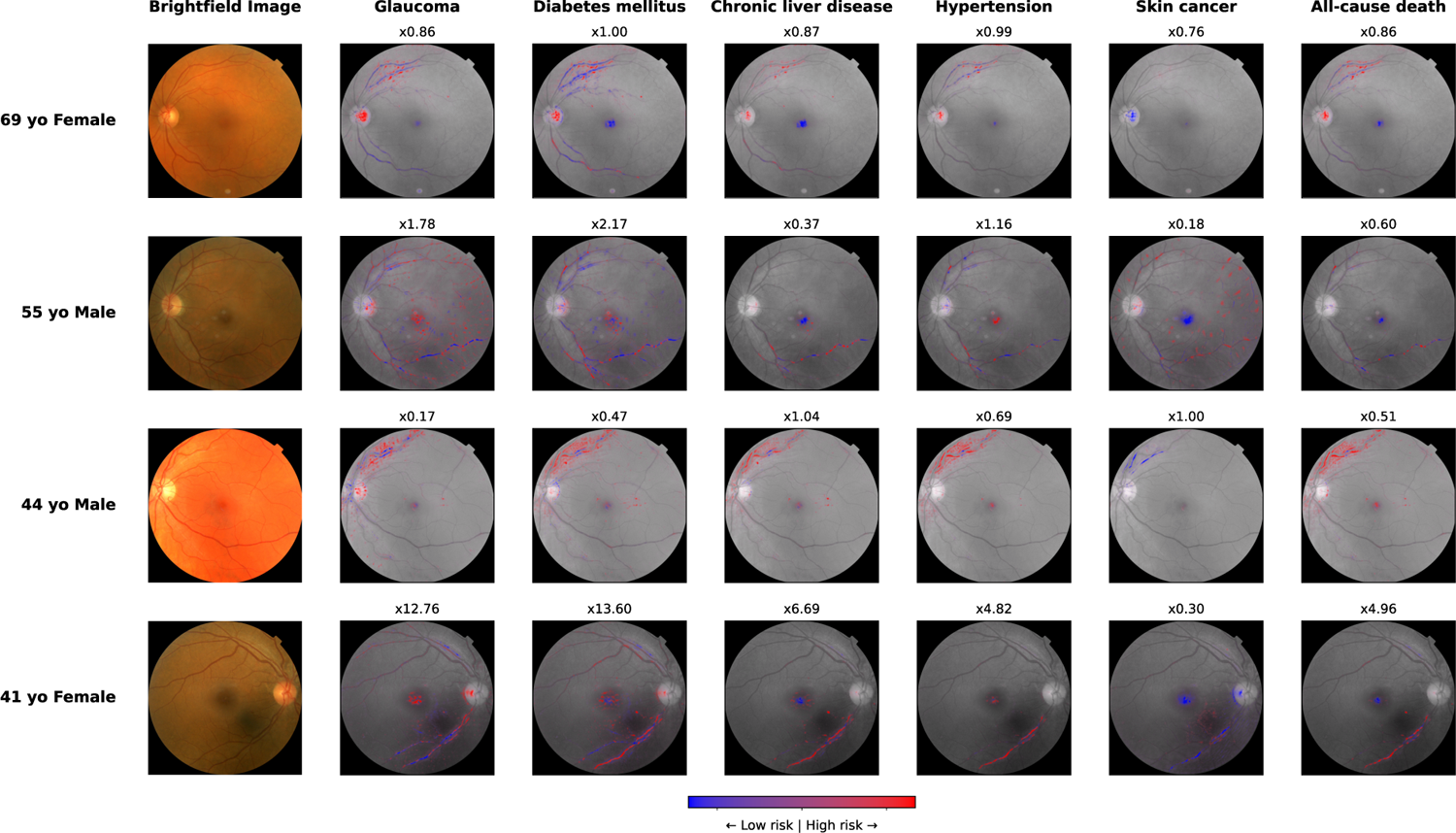
Image attributions characterise the predictive information in the retina. Image attributions for six selected endpoints and four individuals were derived via information bottleneck attribution (IBA) against the retinal state adjusted by age and biological sex. Regions with attributions to increased risk are highlighted in red, while regions with attribution to decreased risk are highlighted in blue. The colour intensity corresponds to the strength of attribution.

### Explainable AI and biological insights from genetic analysis

We next sought to investigate possible biological mechanisms underlying risk estimates across a representative range of 84 diseases across the phenome (n=76 eye unrelated phenotypes). We identified a total of 1385 genetic variant-risk state associations that met genome-wide significance (p<5×10^-8^) distributed across 178 genomic loci, including 35 loci associated with ten or more risk estimates (Figure 5 and Supplementary Table 9, see Methods for details), indicating a broad, and partly shared genetic basis for these risk estimates. Most of the loci (96.0%) have previously been reported in the GWAS catalogue (r^2^>0.1; 62.4% likely the same genetic signal with r^2^>0.8; download: 31/10/2023), demonstrating the biological relevance and plausibility of our findings.

**Figure 5:**
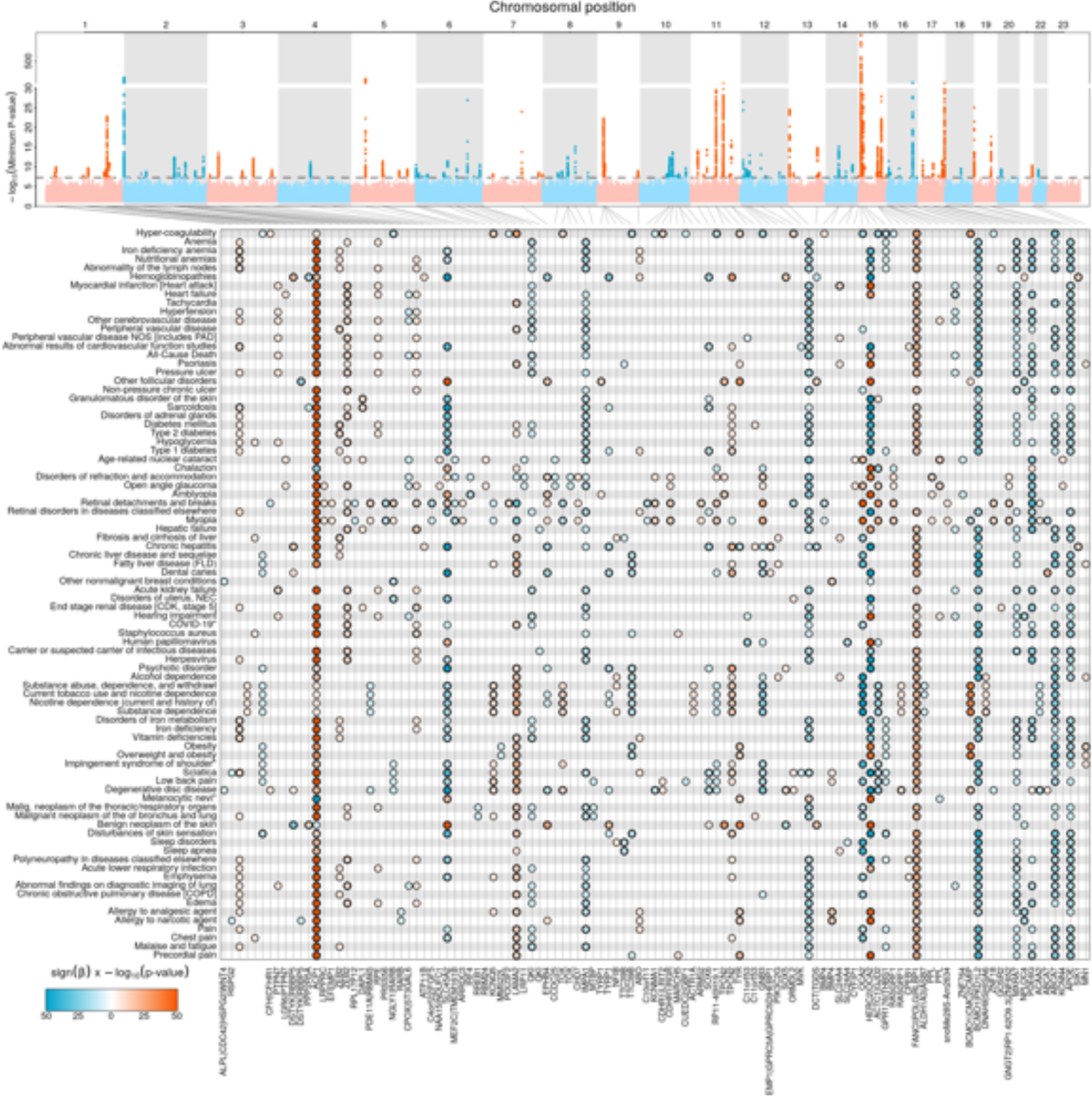
Retinal predictors have a common genetic basis. **a)** Summary Manhattan plot displayed the strongest identified association value (-log_10_(*minimum p-value*)) for each variant over the 84 analysed endpoints. **b)** Individual associations between identified genetic loci and investigated diseases. The black lines indicate the relation between variants and gene loci, positive associations (i.e. increasing the retinal modification of background risk) are coloured in red, while negative associations (i.e. decreasing the retinal modification of background risk) are marked in blue. The colour intensity indicates the estimated effect size.

More than half (n=94) of the independent loci have been previously associated (r^2^>0.1) with morphological and pathological characteristics of the eye, including aspects captured (e.g., a highly pleiotropic missense variant at *ACP1* previously linked to retinal vascular fractal density^42^ and accelerated ageing of the eye^43^) and not captured by image attribution techniques. For example, pleiotropic loci determining eye colour (e.g., rs12913832 mapping to *HERC2-OCA2* associated with, e.g., the risk of hemoglobinopathies: beta=-0.35, p<10^-762^; or rs16891982 mapping to *SLC45A2* associated with, e.g., the risk of chronic hepatitis: beta=-0.35, p<2.8×10^-88^), associating with retinal nerve fibre layer thickness (e.g., rs5442 a missense variant in *GNB3* associated with the risk for degenerative disc disease: beta=-0.11, p<6.74×10^-23^), or age-related macular degeneration (e.g., rs11200634 mapping to *ARMS2* associated with the risk of retinal detachments and breaks: beta=0.05, p<2.26×10^-10^). We also identified genomic regions (e.g., rs11840593 associated with the risk of heart failure: beta=-0.06, p<9.00×10^-22^) that encode multiple genes involved in lens morphology, like *GJA3* involved in the formation of lens fibre gap junctions. The importance of morphological and developmental features of the eye for risk prediction was also apparent from enriched pathways among risk loci, including the development of the ‘melanosome membrane’ (enriched for the risk of hemoglobinopathies, p<1.2×10^-9^), ‘melanin biosynthetic process’ (enriched for the risk of chronic hepatitis, p<1.1×10^-7^), or ‘optic nerve misrouting’ (enriched for the risk of amblyopia, p<2.9×10^-5^).

We also observed a total of 28 loci that provided insights into the aetiology of predicted risk estimates beyond eye morphology by meeting a stringent Bonferroni-corrected genome-wide significance threshold (p<5.9×10^-9^, accounting for 84 tested outcomes) and that have not been previously reported for eye phenotypes (r^2^<0.1). This included known pleiotropic variants, such as allelic variation at *APOE,* and variants that may manifest in the eye without affecting its function. For example, the missense variant rs7185774 (p.Gly129Asp; *PKD1L2*) and closely correlated variants were associated with 61 risk estimates, including psychotic (beta=-0.08, p<2.3×10^-43^) and chronic liver diseases (beta=-0.08, p<3.1×10^-41^). The locus has previously been associated with plasma levels of total bilirubin^44^, and while malfunctioning of the gene product, polycystin 1 like 2, has been mostly linked to polycystic kidney syndrome, increased levels of bilirubin in the eye, called jaundice, is a common symptom of liver disorders. Among the three so far unreported loci, we identified the low-frequent missense variant rs204781 (MAF=2.0%; p.I109V) within *IMPA1* to be linked to a decreased risk state across 48 diverse diseases (n=33 meeting corrected gw-significance), most strongly for iron deficiency anaemia (beta=-0.16, p<7.2×10^-16^). The gene product, inositol monophosphatase 1 (IMPA1), is essential for (myo-)inositol production from myo-inositol mono- or diphosphates in almost all tissues^45^. Given the broad substrate spectrum, one might speculate that IMPA1 is also directly or indirectly involved in the breakdown of myo-inositol 1,2,3-trisphosphate, which in turn is a potent intracellular iron chelator involved in iron homeostasis^46,47^. These results demonstrate how deep learning-based modelling of complex images coupled with risk prediction can reveal novel biological insights not apparent from studying each layer separately.

## Discussion

Cost-efficient identification of people at the highest risk for rare and complex diseases is paramount to mitigate the increasing burden on healthcare systems globally. Here, we provide evidence that retinal fundoscopy, combined with state-of-the-art machine learning, is a potential low-cost strategy to identify people at risk early, especially for eye, metabolic, and mental health diseases. Other disease domains are likely to benefit from different data modalities. We demonstrate that these achievements are driven by obvious features, like vascularization, and less obvious ones, such as lens morphology, by integrating explainable AI techniques with genetic association analysis.

Automatic disease diagnosis and prediction using retinal images and deep learning techniques has recently gained substantial attention through novel foundation models^27^, but a rigorous and systematic assessment of potential clinical utility across the human phenome has been largely lacking. Our findings suggest that retinal fundus photographs contain predictive information for diseases without known retina-disease associations, including COPD, a subset of malignant neoplasms, and mental disorders, such as major depression. This information may result from disease-specific pathological precursors in the retina or mediated through associated demographic predictors^49^. We observed predictive improvements over and above readily available participant characteristics only for a subset of diseases, highlighting both limitations and potential areas of application. For example, we demonstrated superior performance in detecting individuals with chronic liver disease that may translate into the prevention of liver fibrosis and cirrhosis later in life.

Retinal fundus images have been previously suggested for the primary prevention of cardiovascular diseases^10^. However, retinal fundus images did not improve discriminatory performance over the established cardiovascular risk scores SCORE2, ASCVD, or QRISK3 for any investigated cardiovascular endpoints. Consequently, it is unlikely that retinal fundus images will contribute significantly to the primary prevention of cardiovascular disease in the general population. Our findings are consistent with previous studies that suggest retinal fundus photographs serve as an information proxy, albeit with some loss, for basic demographic (e.g. age and sex), behavioural (e.g. smoking and alcohol consumption), and cardiometabolic predictors (e.g. lipid and blood pressure levels)^10,16,43,50^. Although these predictors can be approximated well, the additional information for many endpoints is small. While further improvements to the modelling or the combination of retinal images with other information may show discriminative improvements for the primary prevention of cardiovascular diseases, our study highlights the necessity of evaluating predictive models in the context of established markers and risk scores.

We used Information Bottlenecks for Attribution^41^ to assess the anatomic regions contributing to disease risk. Reassuringly, we found predictive retinal information predominantly originating from retinal vascularisation patterns, the macula, and the fovea, regions that have been previously indicated in many neurological and cardiometabolic diseases^1–3,10^. This could be explained by shared disease processes or risk factors mediated through and observable in retinal fundus scans. Interestingly, individual features with the highest attribution were generally informative for multiple disease endpoints, indicating that pathological alterations in specific regions capture general information relevant to multiple future diseases.

We identified dozens of genomic regions with a clear link to the eye’s morphological, physiological, and aetiological characteristics and associated disorders. This included loci linked to retinal vascularisation (e.g., *ACP1* linked to the onset of myopia^51^), eye colour (e.g., *SLC45A2* linked to the onset of skin cancer^52^), or the architecture of the lens (e.g., *GAJ3* linked to the onset of cataracts^53^) that could explain the superior performance of our model across the phenome compared to previous studies specifically extracting retinal features^54^. These loci may segregate into two categories: 1) ‘horizontally’ pleiotropic loci associated with disease onset and eye features via independent mechanisms (e.g., missense variants in *TPCN2* affect glucagon secretion from the pancreas as well as skin pigmentation^57^), or 2) ‘vertically’ pleiotropic loci, by which the locus acts on eye phenotypes to cause the disease or *vice versa* (e.g., we observed multiple loci associated with both predicted risk and onset of age-related macular degeneration). Notably, we identified a few loci, like *IMAP1* for iron homeostasis, that provided insights into disease aetiologies not seen in bespoke disease GWAS and hence demonstrated the capability of AI-augmented phenotypes to also derive novel biological insights. Our results demonstrate how genetic information can inform and provide tangible insights into otherwise cryptic features extracted by deep learning models and support the validity of the extracted features due to their anchors in known genomic associations.

Despite our study population consisting of over 61,000 individuals and approximately 685,000 person-years of follow-up, it is important to note that this population is healthier and less deprived than the general UK population^58^, suggesting that recalibration and careful reevaluation of the model in the target population may be necessary before clinical use. Furthermore, we were unable to confirm the discriminatory improvements in the EPIC-Norfolk Eye Study for several endpoints, including ischemic heart disease, skin cancer, and all-cause mortality. This is likely due to the smaller sample size, differences in the populations of these cohorts, and potential differences in data acquisition. To thoroughly evaluate the clinical applicability and impact of our approach, further testing in prospective settings is essential. This involves not only assessing the model’s performance in real-time clinical environments but also understanding its socioeconomic implications. Cost-effectiveness analysis will be critical to determine whether the benefits of early disease detection and intervention using retinal fundoscopy outweigh the associated expenses. Furthermore, the generalizability of our findings needs to be confirmed across diverse cohorts, encompassing variations in demographics, genetics, and environmental exposures. This is particularly important to ensure that the model performs equally well across different populations and does not inadvertently introduce or exacerbate existing health disparities. We restricted genetic analyses to people of white European ancestry given the overall very small sample size (N < 1,000) of people of other ancestries in UK Biobank, and replication of our results in people of different ancestries and other marginalized groups will be essential. Furthermore, the ethical implications of employing AI in healthcare, such as issues related to patient consent, data privacy, and the potential for over-reliance on automated systems, need careful consideration.

In conclusion, we demonstrated that retinal fundus images have potential for clinical application beyond their current scope in identifying early risk factors for various diseases, especially in the fields of eye, metabolic, and mental health diseases. We also demonstrated the limited utility of retinal information beyond basic demographic predictors for many diseases across clinical specialties. Our findings indicate that retinal fundus photography could play a role in developing cost-effective screening strategies, aiding in the early detection and management of several diseases. This approach may offer a valuable tool for healthcare systems in mitigating the burden of disease through early intervention.

## Methods

### Data source and endpoint definition

We used data from the UK Biobank cohort, a sample of healthy volunteers from the UK’s general population. Participants were enrolled from 2006 to 2010 in 22 recruitment centres across the United Kingdom; the follow-up is ongoing. The UK Biobank provides retinal fundus photographs recorded at six of the 22 centres for a subset of ∼68,000 individuals. After data filtering, cleaning, and preprocessing (see the section below), we derived the study population of 61,256 individuals, 33,285 women and 27,971 men, aged in median of 58 (IQR 50, 63) years at the time of their baseline assessment. Participants in the UK Biobank cohort are linked to routinely collected records from hospital records (HES, PEDW, and SMR), and death registries (ONS), providing longitudinal information on diagnosis, procedures, and prescriptions for the entire cohort from Scotland, Wales, and England. Endpoints were defined as the set of PheCodes X^59,60^. After the exclusion of rare endpoints (recorded in <= 100 individuals with retinal fundus photographs), 752 PheCodes X endpoints were included in the development of the models. Due to the adult population, congenital, developmental, and neonatal endpoints were excluded. Time-to-event outcomes were extracted for each endpoint, defined by the first occurrence after recruitment in primary care, hospital, or death records. Detailed information on the predictors and endpoints is provided in Supplementary Table 1+2.

The entire study population was utilised for training the model, deriving the retinal risk estimates, and estimating log partial hazards. However, individuals were excluded from endpoint-specific downstream analysis and metric computation if they were diagnosed with a disease (i.e., had a record of the respective phecode prior to baseline) or were generally not eligible for the specific endpoint (e.g., female participants were excluded from the risk estimation for prostate cancer). The study adhered to the TRIPOD (Transparent Reporting of a Multivariable Prediction Model for Individual Prognosis Or Diagnosis) statement for reporting^61^. The completed checklist can be found in the Supplementary Information.

### Preprocessing of retinal fundus photographs

Before model development and training, the corpus of available retinal fundus photographs was cleaned and preprocessed to exclude corrupted and faulty samples. Retinal images usually consist of a central, circular image structure representing the retinal disc, surrounded by dark areas. However, as the retinal fundus images in the UK biobank were automatically taken during optical coherence tomography and not manually curated, the dataset contains faulty images that are heavily underexposed, overexposed, or may even show the eye from the outside. Therefore, we cleaned the dataset by removing images where the retinal disc was not detectable, either because of over- or under-exposition, over- or under-saturation, lack of focus or frame, or due to strong shifts in either of the three colour channels. Specifically, we tested each dataset image for a detectable retinal disc by first converting the image to grayscale and applying the Canny edge detection operator^62^. If the retinal disc was detectable in the grayscale-converted image, all areas outside the retinal disc in the original image were removed (i.e., set to missing values), and the image was trimmed such that the retinal disc touched all four image borders. Next, we manually curated a set of 1,000 valid images with the aid of ophthalmological domain experts and computed distributions (mean and standard deviations) for the three RGB colour channels and the three HSV channels on this curated set. Subsequently, all images of the remaining dataset were compared w.r.t. the means of their RGB and HSV channels to the distribution of the curated samples. All images with a mean outside the two standard deviation interval in any of the six assessed channels defined by the curated reference dataset were excluded from the analysis. The UK Biobank cohort contains ∼175,000 retinal fundus photographs for ∼85,000 participants from both the left and right eyes. Of these images, ∼137,000 items for ∼68,000 participants were measured at baseline, while the remainder of the images were measured five years after baseline. In order to relate the images to the observed outcomes, the analysis was restricted to images recorded at baseline. After applying the preprocessing pipeline, the cleaned dataset contained 113,122 images for 61,256 participants measured at baseline.

### Machine Learning and Downstream analyses

#### Spatial cross-validation

For model development and testing, we split the data set into six spatially separated partitions based on the location of the assessment centre at recruitment, as described previously^63,64^. We analysed the data in six-fold nested cross-validation, setting aside one spatially separated partition as a test set, aggregating the remaining partitions, and randomly selecting 10% of the aggregated data for the validation set. The individual test set (i.e., the spatially separated partition) remained untouched throughout model development within each of the six cross-validation loops. The validation set was used to validate the fitting progress and checkpoint selection. All six obtained models were then evaluated on their respective test sets. We assumed missing data occurred randomly and performed multiple imputations using chained equations with gradient boosting machines^65^. Imputation models were fitted on the training sets and applied to the respective validation and test sets. Continuous variables were standardised; Categorical variables were one-hot encoded.

#### Retinal risk model

The retinal risk model is a vision transformer network simultaneously predicting a scalar estimate of the risk for future incident disease onset, i.e., a retinal state, for 752 endpoints. The retinal risk model consists of two major parts: a large vision transformer network for feature extraction and a smaller head network to reweigh the features and predict the retinal states. The feature extractor is a large vision transformer (ViT-large ^66^), the encoder part of the retinal image foundation model RETFound consisting of 303 million parameters pre-trained in a self-supervised way on a dataset consisting of over 900,000 colour fundus photographs^27^. A custom head network replaces the original decoder part of the RETFound foundation model. The head network consists of three fully connected linear layers, the first with 256 nodes and leaky rectified linear unit activations and the last with 752 nodes and identity activation as out layer. For each endpoint, and thus for each retinal state, an adapted proportional hazard loss^67,68^ is calculated individually, excluding prevalent events for the specific endpoint. The individual losses are averaged and then summed up to derive the final loss of the retinal risk model.

After architecture development, hyperparameter search was run on train and validation splits of partition zero as a random search over a constrained parameter space tuning batch size, initial learning rate, number of nodes in the head network, number of warmup epochs, and the weight decay. The final models were trained with a batch size of 512 for a maximum of 100 epochs using the NAdamW^69^ optimizer with a weight decay of 0.05, a learning rate of 0.0001, a warm-up period of 8 epochs, and early stopping tracking of the performance on each partition’s validation set with a grace period of 20 epochs. The retinal state model was implemented in Python 3.9 using PyTorch 1.7^70^ and PyTorch-lightning 1.4.

During training, the model receives ‘random resized crops’ of the original retinal fundus photographs to present the model with varying feature resolutions. A ‘random resized crop’ is a percentage-based random crop followed by a resize to a given pixel shape. During training, the model receives random resized crops with crop ratios drawn for each sample from a uniform distribution between 0.08 and 1. The cropped images are subsequently fed through the extensive augmentation pipeline also used for the RETfound foundation model consisting of random resized crops to a size of 224×224 pixels, random horizontal flips with probability 0.5, a RandAugmentaion policy, normalisation, and RandomErasing. All augmentations were implemented with the respective augmentation methods offered in torchvision.transforms as part of the PyTorch Python package^70^ and the PyTorch Image Models (timm) library.

At test time, the predictions on the test sets were generated using test-time augmentation of 100 samples per image with random crop ratios between 0.08 and 1, followed by a centre crop of 224×224 pixels. Thus, each image was fed 100 times through the augmentation pipeline and the network at test time, retrieving 100 predictions. For each endpoint and image, the arithmetic mean of these 100 predictions was used as the final retinal state for this endpoint.

#### Cox proportional hazards models

Cox proportional hazards^30^ (CPH) models were fitted to derive absolute risk predictions from the endpoint-specific retinal risk estimates for the individual endpoints. For each endpoint, we developed models with distinct covariate sets. For all endpoints, we investigated age, biological sex, and the retinal risk state. Additionally, for cardiovascular endpoints, we investigated predictors from established and guideline-recommended scores for the primary prevention of cardiovascular diseases, the SCORE2^35^, ASCVD^36^, and QRISK3^37^. Models were developed in six-fold spatial cross-validation, aggregating all test set predictions for downstream analyses. Harell’s C-Index was calculated with the lifelines package^71^ by bootstrapping both the aggregated test set and individual assessment centres. Statistical inferences about model differences were based on the distribution of bootstrapped differences in the C-Index; models were considered different whenever the 95% CI of the difference did not overlap cross zero. CPH models were fitted with the CoxPHFitter from the Python package lifelines^71^ with default parameters and a step size of 0.5, 0.1, or 0.01 to facilitate model convergence.

#### Bootstrapping

To provide reliable metrics and significance measures, we bootstrapped metrics distributions. For each iteration, metrics were computed on a randomly sampled set of individuals (with replacement). For endpoints with a low number of observed events, in each iteration, sampling was repeated if no observed events were in the subset of drawn individuals. Generally, bootstrapping was applied with 100 iterations if not indicated otherwise.

### Independent validation in the EPIC-Norfolk eye study

To assess the generalizability and robustness of our retinal risk model beyond the initial UK Biobank cohort, we performed external validation using data from the EPIC-Norfolk eye study. This cohort includes 7,248 individuals who underwent fundal photography and were tracked for a range of health outcomes in middle and later life. The validation process was performed on a single compute node within the UCL cluster. For this analysis, endpoints were extracted from Hospital Episode Statistics (HES) and Office for National Statistics (ONS) records up to the censoring date of June 17, 2019. Only endpoints with a minimum of 100 incident events were considered to ensure statistical reliability. The Retinal Risk model, initially trained on UK Biobank data, was directly applied to the EPIC-Norfolk study without further training. Baseline feature preprocessing and imputation followed identical procedures as established in the UK Biobank analysis. However, images labeled as of “poor” quality were excluded from the analysis. Evaluation of model efficacy mirrored the procedures implemented for the UK Biobank, involving 100 bootstrap samples to provide reliable metrics and significance measures.

### Genetic analyses

#### Genotyping, quality control, and participant selection

Details on genotyping for UKBB have been reported in detail by Bycroft et al.^72^. Briefly, we used data from the ‘v3’ release of UKBB containing the full set of Haplotype Reference Consortium (HRC) and 1000 Genomes imputed variants. We applied recommended sample exclusions by UKBB, including low-quality control values, sex mismatch, and heterozygosity outliers. We defined a subset of ‘white European’ ancestry by clustering participants based on the first four genetic principal components derived from the genotyped data using a k-means clustering approach with k=5. We classified all participants who belonged to the largest cluster and self-identified as being ‘white,’ ‘British’, ‘Any other white background’, or ‘Irish’ as ‘white European’. After applying quality control criteria and dropping participants who had withdrawn their consent, 444,441 UKBB participants were available for analysis with genotype data. We then merged genetic data with predicted risk estimates based on per PheCode exclusions, resulting in varying numbers for each analysed risk state (n=39,930-51,882).

We used only called or imputed genotypes and short insertions/deletions (here commonly referred to as SNPs for simplicity) with a minor allele frequency (MAF)>1.0%, imputation score >0.4, and within Hardy-Weinberg equilibrium (pHWE>10^-15^). This left us with 10,027,927 autosomal and X-chromosomal variants for statistical analysis. GRCh37 was used as reference genome assembly.

#### Genome-wide association studies

We performed genome-wide association studies (GWAS) for 84 risk estimates (Supplementary Table 9) using REGENIE v2.2.4 via a two-step procedure to account for population structure as described in detail elsewhere^73^. We used a set of high-quality genotyped variants (MAF>1%, MAC>100, missingness <10%, pHWE>10^-15^) in the first step for individual trait predictions using the leave one chromosome out (LOCO) scheme. These predictions were used in the second step as offset to run linear regression models. Each model was adjusted for age, sex, genotyping batch, assessment centre, and the first ten principal genetic components. We excluded participants with a record of the respective phecode at baseline from genetic analysis. We used a standard genome-wide significance threshold to declare significant findings but report specifically how many of the findings also met a more stringent threshold, further corrected for the number of outcomes considered (p<1.1×10^-9^). We used regional clumping to select approximately independent genetic findings by first defining a 1Mb window around regional sentinels and afterwards merging closely related regions. We treated the extended MHC region as one (chr6:25.5-34.0Mb). We used LD-score regression to test for genomic inflation and calculated the SNP-based heritability (LDSC v1.0.1)^74^.

#### Candidate causal gene assignment

We used four resources to provide candidate causal genes at each loci identified.

1. Statistical colocalisation with gene expression (cis-eQTL) in 49 tissues from the GTEx project (v8)^75^. Briefly, we considered all protein-coding genes or processed transcripts encoded in a 1Mb window around risk loci and performed statistical colocalization^76^ to test for a shared genetic signal between a phecode and gene expression. We adopted a recently recommended prior setting^77^ with p12=5×10-6.
2. Statistical colocalisation with plasma protein levels (cis-pQTL) from our previous study^72^ implemented in the same way as for eQTL.
3. Annotation of lead genetic variants and potential functional proxies (r2>0.6) using the Variant Effect Predictor software (v.106). We considered all genetic variants with a score of twelve or lower as’ functional’ based on the VEP recommendations.
4. Nearest gene based on the gene body or transcription start site.

For each candidate gene at a locus, we computed weighted score across all six criteria, thereby colocalisation evidence was divided based on moderate (50%>PP<80%; 0.5 points) or high (PP>80%; 1 point) confidence and functional variants were assigned highest priority (2 points). We reported gene(s) with the highest score as candidate causal gene(s) at each locus.

#### Calculation of image attributions

To determine which image regions most contribute to a prediction on an individual level, we calculated information bottleneck attributions (IBA)^41^ for a selection of six endpoints with significant added predictive information over basic demographic predictors. Information bottleneck attribution (IBA) allows the identification of important image regions by inserting an information bottleneck at lower levels of a neural network, learning a parameter mask to retain predictive information while omitting irrelevant information. Individuals for the attribution maps were selected from the union set of the top 10 individuals with the highest risk modification per endpoint. In order to generate attributions for an image, IBA is applied for 512 crops with a crop ratio of 0.5 and images are subsequently aggregated to retrieve a full retinal fundus photograph. Specifically, using the described procedure, we calculate attributions with two information bottlenecks optimised to maximise information increasing or decreasing the predicted retinal state (i.e., the log partial hazards quantified by the adapted Cox PH loss^67,68^), resulting in two independent attribution maps. For visualisation, we calculate the direction of the aggregated attribution (represented by the colour map) as the difference between the positive (risk-contributing) and negative (risk-reducing) attributions and the strength of the attribution as the maximum of the absolutes of both attribution maps (represented by the alpha variable in the visualisation). To reduce border attribution effects and highlight all non-marginal features even more, attribution maps were masked circularly by the 16 outmost pixels in each direction. To create image attributions adjusted for biological sex and age, we trained another retinal risk model with the same convolutional network architecture. However, in this model, we added biological sex and age as supplementary features to the custom head network. By incorporating the demographic features at the input stage of the head network and the information bottlenecks at the early stages of the convolutional network, this approach ensured that the convolutional network extracted only the information that was additive to the demographic features.

## Code availability

The code is publicly available in the following Github repositories: Deep learning model training (https://github.com/thbuerg/RetinalRisk), Evaluation in UK Biobank (https://github.com/lukl95/22_retina_phewas_evaluation), Evaluation in EPIC (https://github.com/JakobSteinfeldt/23_retina_epic).

## Data Availability

UK Biobank data, including retinal fundus photographs, are available to bona fide researchers upon application at http://www.ukbiobank.ac.uk/using-the-resource/. Detailed information on predictors and endpoints used in this study is presented in Supplementary Tables 2-3. EPIC-Norfolk data are available for the scientific community, and researchers are invited to apply for data access at https://www.epic-norfolk.org.uk/for-researchers/data-sharing/data-requests/.

## Acknowledgements

This research was conducted using data from UK Biobank, a major biomedical database (https://www.ukbiobank.ac.uk/) via application no. 51157 and 44448. We are grateful to all the participants and all members of the study teams who have enabled this research. Further, we used data from the EPIC-Norfolk study via the application of APK. The EPIC-Norfolk study (DOI 10.22025/2019.10.105.00004) has received funding from the Medical Research Council (MR/N003284/1 and MC-UU_12015/1) and Cancer Research UK (C864/A14136). Both studies have received ethical approval from their respective institutional review boards and have obtained informed consent from participants. We are grateful to all the participants who have been part of the project and to Nicholas Wareham and the many members of the study teams at the University of Cambridge who have enabled this research. This project has been funded by the Charite - Universitaetsmedizin Berlin and the Einstein Foundation Berlin, through the Einstein BIH Visiting Fellowship awarded to J.D. The study has been supported by the BMBF-funded Medical Informatics Initiative (HiGHmed, 01ZZ1802A - 01ZZ1802Z) and the Deutsche Forschungsgemeinschaft (DFG, German Research Foundation) Project-ID 437531118 SFB 1470. APK is supported by a UK Research and Innovation Future Leaders Fellowship, an Alcon Research Institute Young Investigator Award and a Lister Institute for Preventive Medicine Award. This research was supported by the NIHR Biomedical Research Centre (BRC) at Moorfields Eye Hospital and the UCL Institute of Ophthalmology. PJF and APK received salary support from the NIHR BRC at Moorfields Eye Hospital. PJF received an unrestricted grant from the Alcon Research Institute, and support from the Richard Desmond Charitable Trust.

## Author contributions

T.B., L.L., J.S., M.P., B.W. and R.E. conceived and designed the project. T.B., L.L., J.S., M.P. and B.W. implemented models, conducted experiments, and performed data analysis. L.H., S.K., H.S. supported the analysis. A.P.K and R.L. supported us in the external validation. A.P.K, J.U.z.B., R.L., P.J.F. and C.L. provided methodological support and contributed to the discussion of the results. LH, SEK, OZ, AMJ provided clinical ophthalmological expertise. T.B., L.L., J.S., M.P., B.W. and R.E. wrote and prepared the manuscript. All authors read, revised, and approved the manuscript.

## Competing interests

U.L. received grants from Bayer, Novartis, Amgen, consulting fees from Bayer, Sanofi, Amgen, Novartis, Daichy Sankyo, and honoraria from Novartis, Sanofi, Bayer, Amgen, Daichy Sankyo. J.D. received consulting fees from GENinCode UK Ltd, honoraria from Amgen, Boehringer Ingelheim, Merck, Pfizer, Aegerion, Novartis, Sanofi, Takeda, Novo Nordisk, Bayer, and is chief medical advisor to Our Future Health. R.E. received honoraria from Sanofi and consulting fees from Boehringer Ingelheim. APK has acted as a paid consultant or lecturer to Abbvie, Aerie, Allergan, Google Health, Heidelberg Engineering, Novartis, Reichert, Santen, Thea and Topcon. PJF received personal fees from Allergan, Carl Zeiss, Google/DeepMind and Santen, outside the submitted work. All other authors do declare no competing interests. O.Z. received grants from Bayer, Boehringer-Ingelheim, Novartis, consulting fees from Allergan/AbbVie, Bayer, Boehringer-Ingelheim, Novartis, Omeicos, Oxular, Roche, SamChungDang Pharma, and honoraria from Allergan/AbbVie, Bayer, Boehringer-Ingelheim, Novartis, Roche.

### Tables

**Table 1:** The study population.

## Supplementary Figures

**Supplementary Figure 1:**
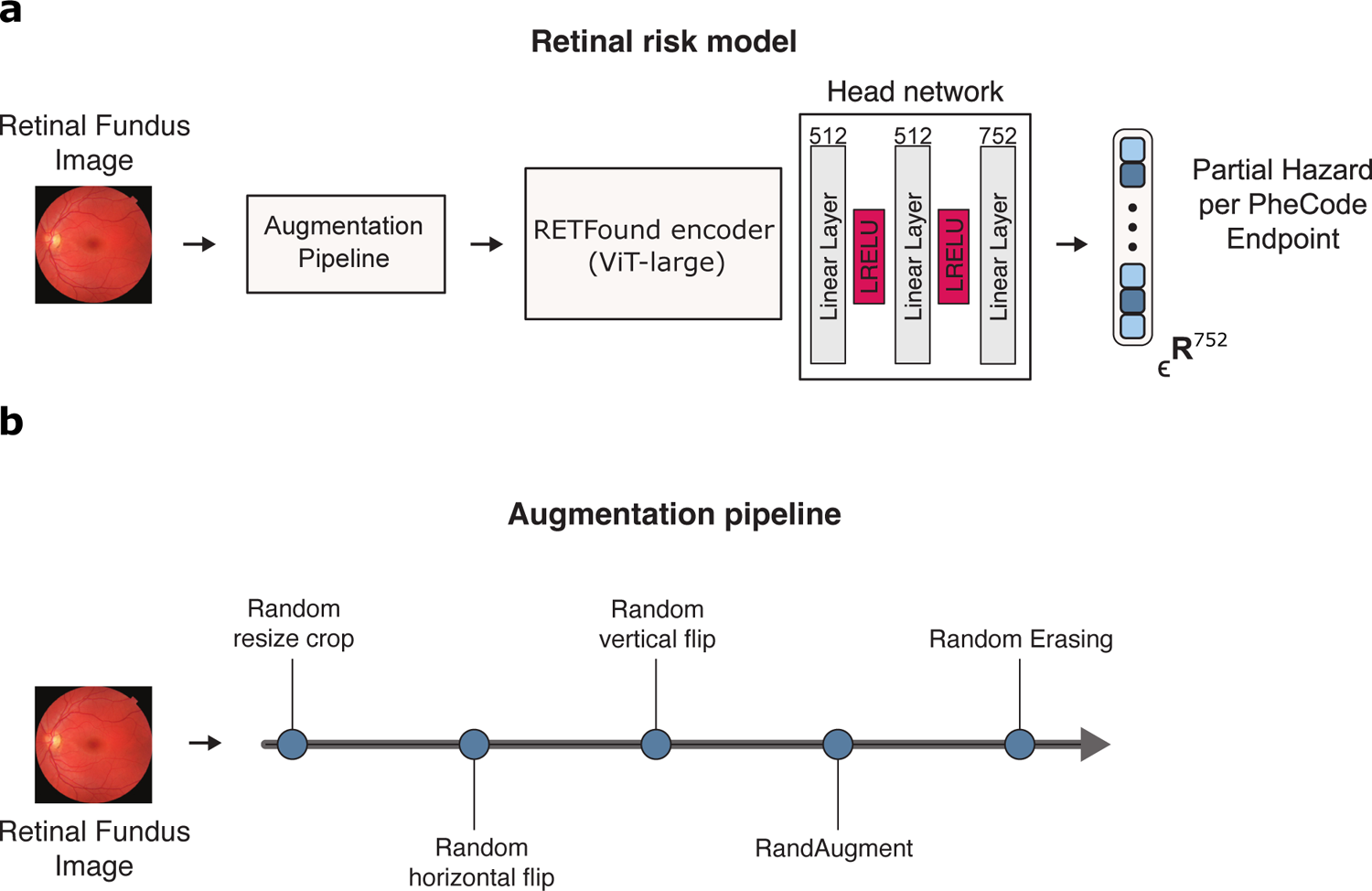
Architecture of the retinal risk model and the augmentation pipeline. **a)** The retinal risk model is a vision transformer encoder architecture from the RETFound fundus photograph model, with a custom head network predicting log partial hazards for 752 endpoints simultaneously from a 224×224-sized image input. **b)** For training, retinal fundus images are pushed through the augmentation pipeline sequentially, applying each augmentation with a random probability before being fed to the retinal risk model.

**Supplementary Figure 2:**
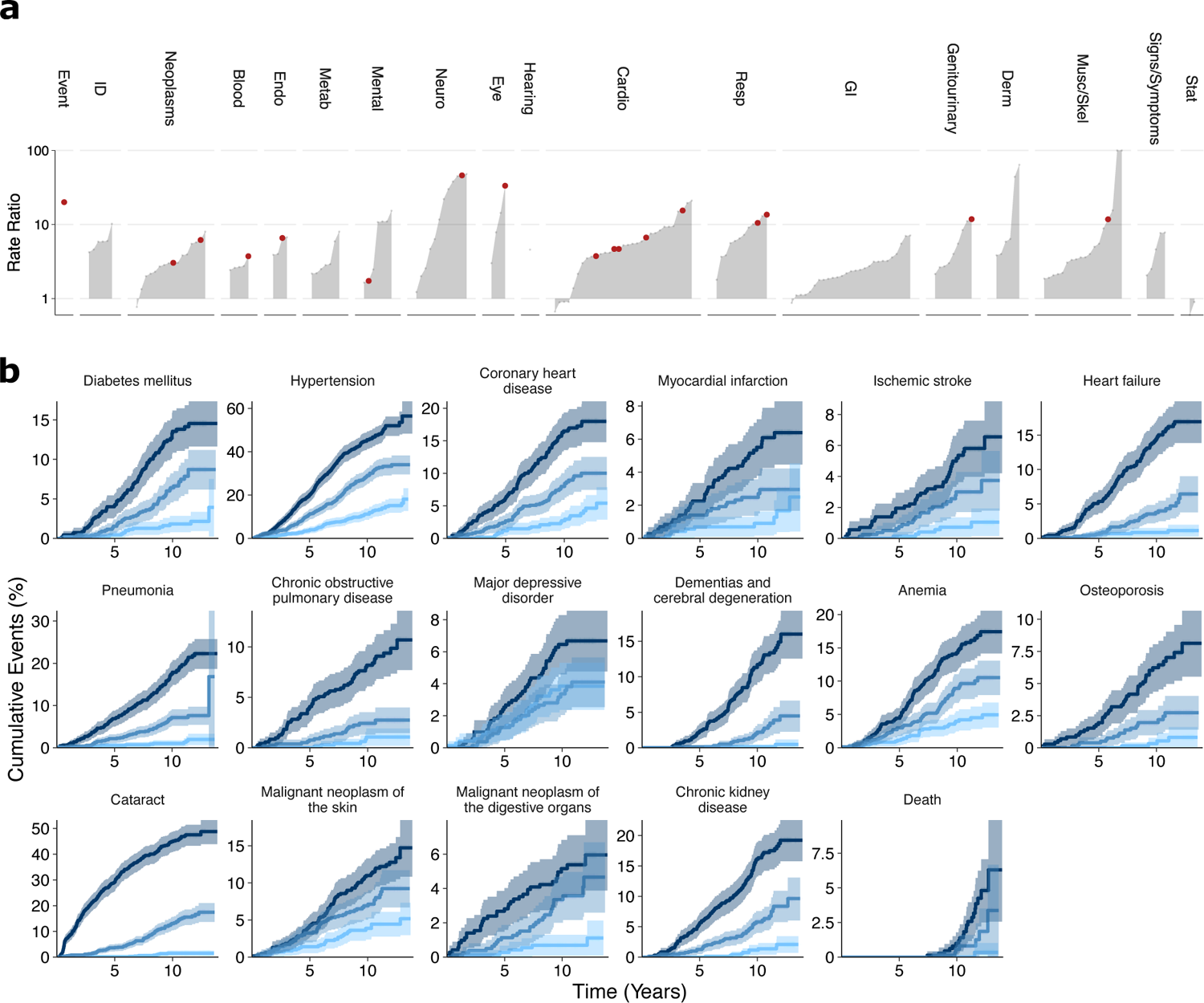
Stratification in external validation in the EPIC-Norfolk Eye Study. **a)** Ratio of incident events in the Top 10% compared with the Bottom 10% of the estimated risk estimates for 173 endpoints (>= 100 incident events). Red dots indicate the selected endpoints displayed in **b)** Cumulative event rates for a selection of endpoints for the Top 10%, median, and Bottom 10% of risk percentiles over 15 years. Statistical measures were derived from 7,248 individuals. Individuals with prevalent diseases were excluded from the endpoints-specific analysis.

**Supplementary Figure 3:**
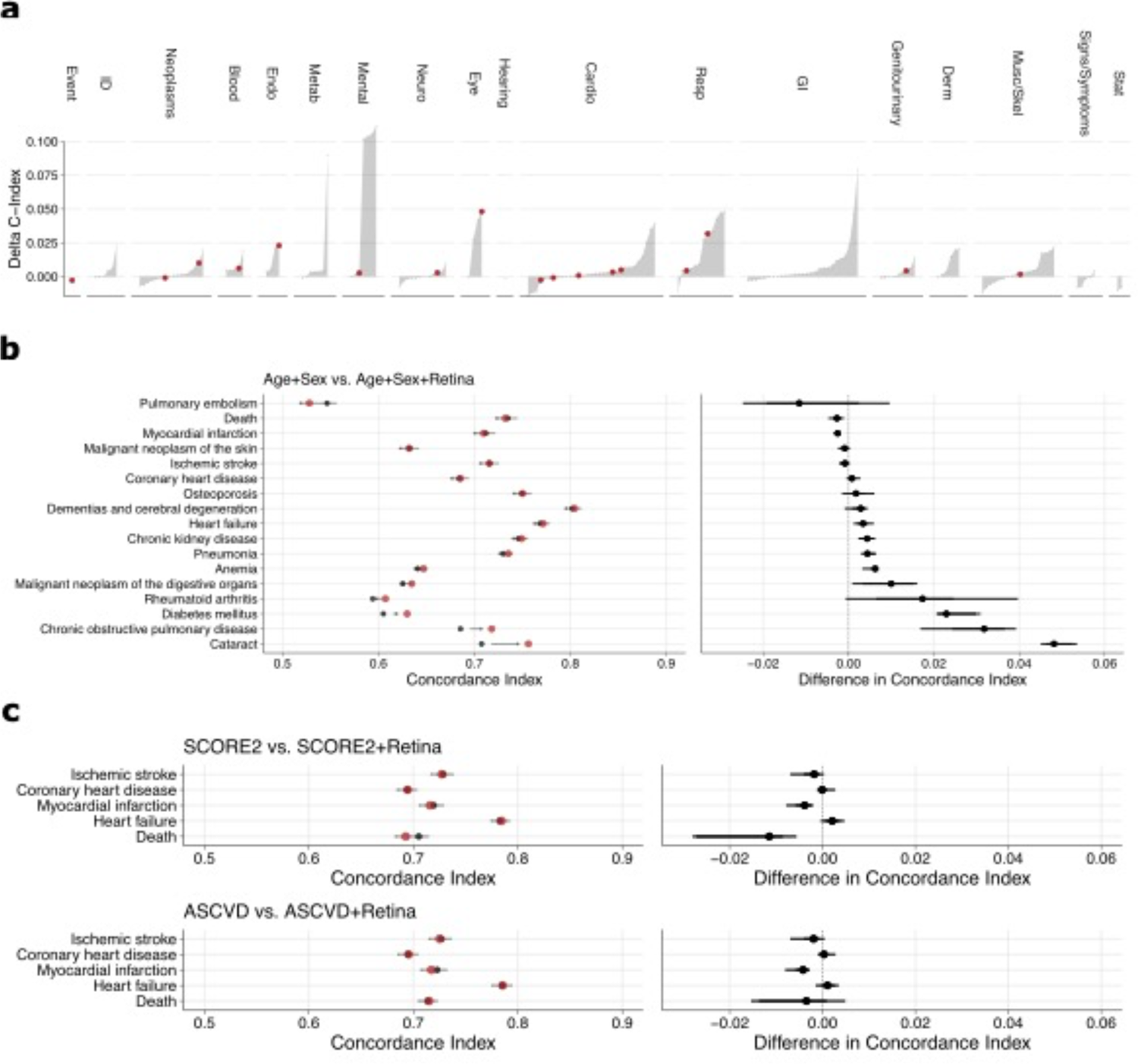
Discrimination in external validation in the EPIC-Norfolk Eye Study. **a)** Differences in discriminatory performance quantified by the C-Index between CPH models trained on Age+Sex and Age+Sex+Retina for 173 endpoints. Significant improvements over the baseline model (Age+Sex, age, and biological sex only) are found for 55 (32%) of the 173 investigated endpoints. Red dots indicate selected endpoints in **b)** Absolute discriminatory performance in terms of C-Index comparing the baseline (Age+Sex, black point) with the added retinal information (Age+Sex+Retina, red point) for selected endpoints. **c)** Discriminatory performances (left) and direct differences (right) in terms of C-index between CPH models trained on sets of established cardiovascular predictors (SCORE2 and ASCVD as black dots) and a simplified risk model based on Age+Sex+Retina (red dots). Dots indicate medians and whiskers extend to the 95% confidence interval for a distribution bootstrapped over 100 iterations.

**Supplementary Figure 4:**
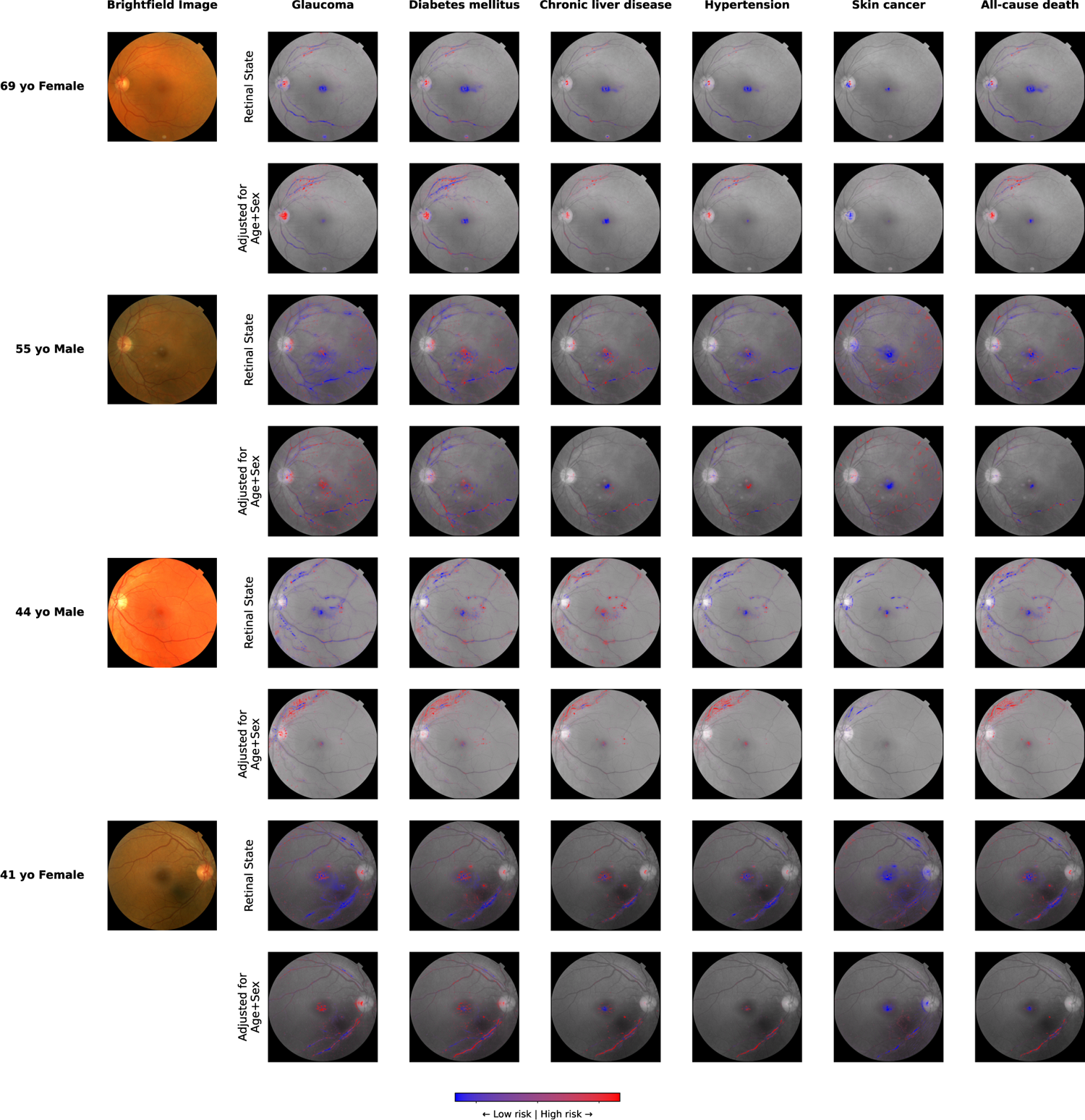
Comparison of adjusted and unadjusted attributions Image attributions for five selected endpoints and four individuals were derived via information bottleneck attribution (IBA) against the retinal state (top row) and the retinal state adjusted by age and biological sex (bottom row). Regions with attributions to increased risk are highlighted in red, while regions with attribution to decreased risk are highlighted in blue. The colour intensity corresponds to the strength of attribution.

